# Reduction of exposure to simulated respiratory aerosols using ventilation, physical distancing, and universal masking

**DOI:** 10.1101/2021.09.16.21263702

**Authors:** Jayme P. Coyle, Raymond C. Derk, William G. Lindsley, Theresa Boots, Francoise M. Blachere, Jeffrey S. Reynolds, Walter G. McKinney, Erik W. Sinsel, Angela R. Lemons, Donald H. Beezhold, John D. Noti

**Affiliations:** Health Effects Laboratory Division, National Institute for Occupational Safety and Health, Centers for Disease Control and Prevention, Morgantown, West Virginia, USA

**Keywords:** Exposure reduction, ventilation, universal masking, physical distancing

## Abstract

To limit community spread of SARS-CoV-2, CDC recommends universal masking indoors, maintaining 1.8 m of physical distancing, adequate ventilation, and avoiding crowded indoor spaces. Several studies have examined the independent influence of each control strategy in mitigating transmission in isolation, yet controls are often implemented concomitantly within an indoor environment. To address the influence of physical distancing, universal masking, and ventilation on very fine respiratory droplets and aerosol particle exposure, a simulator that coughed and exhaled aerosols (the source) and a second breathing simulator (the recipient) were placed in an exposure chamber. When controlling for the other two mitigation strategies, universal masking with 3-ply cotton masks reduced exposure to 0.3–3 µm coughed and exhaled aerosol particles by > 77% compared to unmasked tests, whereas physical distancing (0.9 or 1.8 m) significantly changed exposure to cough but not exhaled aerosols. The effectiveness of ventilation depended upon the respiratory activity, i.e., coughing or breathing, as well as the duration of exposure time. Our results demonstrate that a combination of administrative and engineering controls can reduce personal inhalation exposure to potentially infectious very fine respiratory droplets and aerosol particles within an indoor environment.

**PRACTICAL IMPLICATIONS:**

- Universal masking provided the most effective strategy in reducing inhalational exposure to simulated aerosols.
- Physical distancing provided limited reductions in exposure to small aerosol particles.
- Ventilation promotes air mixing in addition to aerosol removal, thus altering the exposure profile to individuals.
- A combination of mitigation strategies can effectively reduce exposure to potentially infectious aerosols.

## 1 INTRODUCTION

The association between human respiratory infection transmission by respiratory droplets and aerosols is well-established for several known pathogens.^1^ Given that the average individual spends > 90% of their day indoors^2^, there has been intense focus on factors associated with indoor transmission of SARS-CoV-2, the virus that causes COVID-19.^3,4^ Epidemiological investigations highlight the role of congested, poorly ventilated spaces with high levels of secondary attack rates and community transmission.^5,6^ While the specific contribution of respiratory droplets and aerosols remains a topic of active research, increasing evidence of asymptomatic and pre-symptomatic individuals^7^ contributing to community COVID-19 transmission suggests that very fine respiratory droplets and aerosol particles can spread SARS-CoV-2.^8^ To minimize exposure risks, the Centers for Disease Control and Prevention (CDC) recommends several mitigation strategies to limit COVID-19 transmission, including wearing masks, maintaining physical distances, and avoiding crowded indoor and outdoor spaces, among other strategies.^9,10^

Universal masking reduces respiratory aerosol exposure through source control, i.e., limiting the release of infectious very fine respiratory droplets and aerosol particles into the ambient environment at the point of generation. While the generalized effectiveness of masking for source control has been established,^11-13^ its effectiveness is neither absolute nor uniform in practice. Variations in filtration efficiency, air flow resistance, user compliance, and mask fit can limit the effectiveness of masks as source control and protection for the wearer.^14,15^ Despite these limitations, comparison of COVID-19 cases among states employing mask mandates demonstrate an association between universal masking and reduced incidence rates^16-18^ as well as community transmission of COVID-19.^19,20^

Physical distancing reduces infectious material transfer via respiratory-derived droplets and aerosol particles. Routine respiratory actions, such as breathing and normal speech, produce micron and sub-micron scale particles that can remain airborne for minutes to hours.^21^ By comparison, coughing, loud speech, and singing can project aerosols and droplets over greater distances, thus potentially increasing the probability for pathogen transmission. For example, droplets and aerosols produced by coughing may travel up to 8 m.^22^ Analysis of a super spreader event among a cohort of choir members, all of whom were unmasked and within 1.8 m of physical distance during practice, estimated the SARS-CoV-2 attack rate as between 53.3% to 86.7%.^23^ Correlative analyses support an association between physical distancing policies and reduction in COVID-19 incidence,^24,25^ and agree with case-control studies.^26^

Engineering controls, such as room ventilation, are an effective and reliable strategy to ensure good air quality while mitigating infection transmission in the indoor environment.^8,27^ While evidence clearly shows increasing ventilation rates as an effective measure in exposure mitigation,^28,29^ air flow patterns can influence the dispersion of potentially infectious respiratory aerosols and personal exposure,^30^ particularly in confined spaces.^31^ The overall effectiveness of ventilation can be difficult to generalize since ventilation is unique to each room and operates alongside other exposure mitigating strategies, such as masking and physical distancing. As such, the current investigation examines the combined effect of physical distancing, universal masking, and ventilation on exposure to simulated very fine respiratory droplets and aerosol particles generated during breathing and coughing within a controlled indoor environment. The results of this investigation quantitatively examine the contribution of the matrix of controls employed on respiratory infection mitigation strategies within the indoor environment.

## 2 MATERIALS AND METHODS

### 2.1 Environmental Chamber and Ventilation

The testing environment consisted of an environment chamber measuring 3.15 m × 3.15 m × 2.26 m (gross internal volume of 23.8 m^3^, Figure 1). An internal re-circulating high-efficiency particulate air (HEPA) filtration system (Flow Sciences, Inc.; Leland, NC) was used to reduce background aerosol/particle concentrations to near-zero prior to each experiment. The HEPA system consisted of a 10.8 cm inlet duct positioned along the left wall 55.9 cm from the ground leading to the central motor/filter unit and an outlet duct positioned along the right wall at a height of 2.19 m from the ground; no external fresh air was introduced into the environmental chamber during experimentation. Six Grimm 1.108 optical particle counters (OPCs; GRIMM Aerosol Technik Ainring GmbH & Co. KG; Ainring, Germany) were positioned at a height of 152 cm throughout the chamber. The OPCs measured particle concentrations in channels ranging from 0.3 to 3.0 µm at a frequency of 1 Hz, except for one OPC sampler at 0.167 Hz. Four OPCs were affixed to telescopic stands 152 cm above the floor and referred to as “area samplers.” One OPC was positioned 3.2 cm next to the mouth central axis and anteriorly planar to the mouth opening of the recipient simulator (see below) and fit behind a mask affixed to the simulator; this position is denoted as “at the mouth of the breather” for presentation purposes. The remaining OPC was positioned 8.9 cm next to the mouth central axis and anteriorly planar to the mouth opening of the recipient simulator to allow for measurement in the personal breathing zone outside of a mask affixed to the simulator. All OPCs were controlled and data logged using a custom program in LabVIEW v. 2009 (National Instruments; Austin, TX).

**Figure 1.**
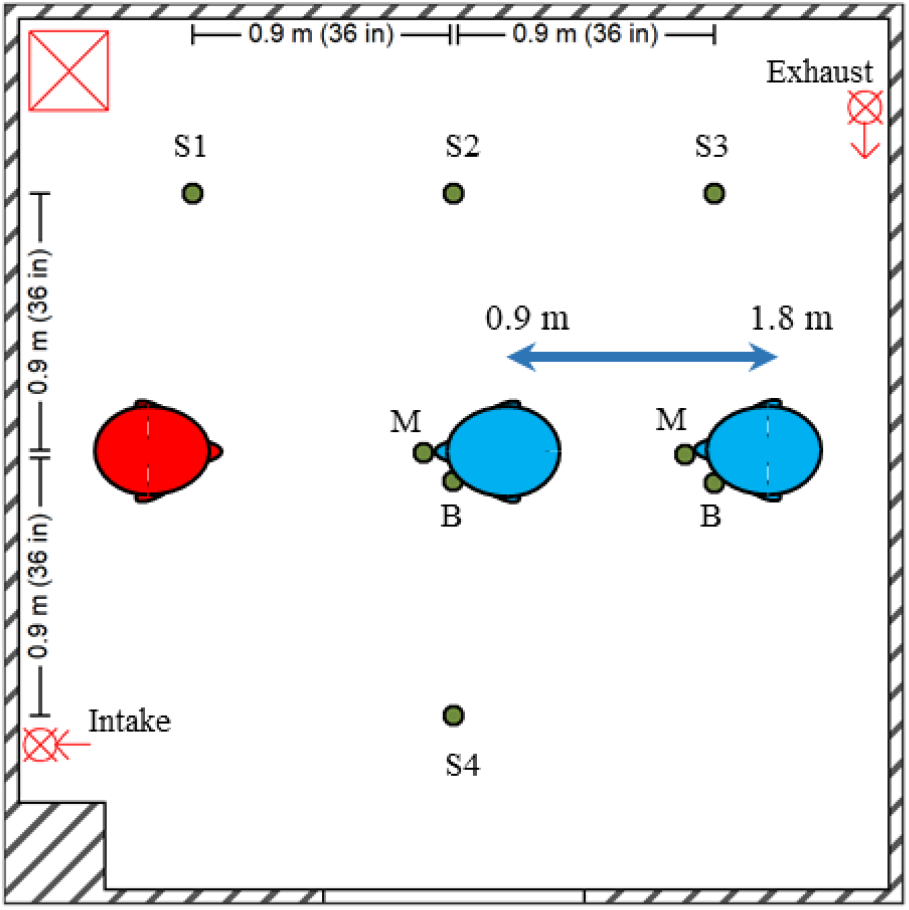
Experimental Setup and Simulators. Diagram of environmental chamber setup showing positions of the aerosol source simulator (red), recipient simulators (blue; position adjustable between 0.9 m and 1.8 m), and optical particle counters (green dots) for area measurements (S1– 4) and personal breathing zone measurements at the mouth (M) and beside the head (B) of the recipient. The HEPA system intake and exhaust are shown with the HEPA filter and blower unit demarcated by the red square containing an “X”.

In addition to particle removal, the HEPA system provided ventilation, with a variable transformer (Staco Energy Products, Co.; Miamisburg, OH) used to set the HEPA system flow rate. Air exchange rates were determined via single-point measurement of the linear air flow at the inlet duct using a Model 5725 VelociCalc rotating vane anemometer (TSI, Inc.; ; Shoreview, MN) equipped with a tapered air cone (TSI, Inc.). The inlet duct was straightened for a length of > 10 diameters from the inlet to minimize turbulent flow during anemometer readings for air changes per hour (ACH) derivation. The HEPA system was set to 0 ACH, 4 ACH (15.3 m^3^/min flow), 6 ACH (22.9 m^3^/min), and 12 ACH (45.9 m^3^/min); calculations assumed zero leakage into the chamber. Effective air filtration rates were derived empirically. Briefly, the chamber was saturated with particles using a stand-alone TSI Model 8026 generator until the 0.3–0.4 um particle size channel reached 10^5^ particles per liter under constant mixing using a household fan. The particles for effective air changes per hour were generated using a 1% solution of NaCl in distilled water formulated from 100 mg tablets provided with the TSI Model 8026 generator as per manufacturer’s instructions. After a 15-minute air settling period, the HEPA filtration system was set to the desired ACH based on anemometer measurements. Particle concentrations were measured for 20 minutes using five of the six OPCs to derive particle exponential decay curves spatially throughout the chamber. Theoretical particle exponential decay curves were modeled from the three smallest size bins (0.3–0.4 µm, 0.4–0.5 µm, and 0.5–0.65 µm) assuming negligible loss to chamber surfaces and aerosol agglomeration using MATLAB v. 9.6 (Mathworks; Natick, MA). The slope of the modeled particle decay was assumed to be first order as per equation:

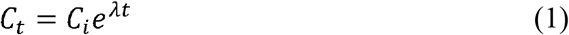

Where:

*C*_*t*_ is the particle concentration at time t (#/cm^3^)

*C*_*i*_ is the initial particle concentration at time zero (#/cm^3^)

*e* is Euler’s number, approximated to 2.71828

*λ* is the slope of particle concentration change over the time (#/cm^3^/second)

*t* is time (seconds)

Empirical concentrations of particles measured by the five area OPCs were then fitted via loglinear regression and the resultant decay coefficient (λ) derived to estimate the effective OPC-specific ACH.

### 2.2 Aerosol Source and Simulators

The source simulator had a head form with pliable skin (Hanson Robotics; Plano, TX) as described in previous work.^12^ For these tests, a single cough and two versions of simulated breathing were examined. The simulated very fine respiratory droplets and aerosol particles (herein designated as aerosol) were produced with a 14% w/v KCl solution nebulized by a single jet Collison atomizer (BGI, Inc.; Butler, NJ) with an inlet pressure of 103 kPa (15 lbs./in^2^) prior to passive drying (Model 3062; TSI, Inc.), dilution with dry filtered air at 10 l/min (single cough tests) or 15 l/min (breathing tests), and neutralization by an ionizer (Model HPX-1, Electrostatics, Inc.; Hatfield, PA). The coughing modality was performed by loading the simulator elastomeric bellows with test aerosol, followed by a single 4.2 l rapid exhalation at a peak flow rate of 11 l/min;^31^ the simulator did not breathe following the cough. For breathing tests, the simulator breathing rate was 12 breaths/min with a tidal volume of 1.25 l and ventilation rate of 15 l/min. The breathing parameters correspond to the ISO standard for females performing light work.^32^ For the breathing modality, the nebulizer was cycled 10 seconds on and 50 seconds off continuously throughout the test duration. Tests were conducted for a duration of 15 minutes, except for a limited subset of testing conditions which were conducted for 60 minutes. As an additional examination of the time-dependency of ventilation in reducing recipient exposure, additional tests were conducted using a modified aerosol generation cadence during the breathing action. During these tests, the nebulizer generated aerosol continuously for the first 3 minutes of the test, after which the nebulizer was turned off, and are henceforth designated short-term aerosol generation tests.

To simulate source aerosol exposure to a recipient, a breathing simulator (Warwick Technologies Ltd.; Warwick, UK) with a pliable skin head form (Respirator Testing Head Form 1 – Static; Crawley Creatures Ltd.; Buckingham, UK) was placed upon a mobile cart to enable alteration of the distance between source and the recipient. The mouth of the recipient simulator head form was positioned 152 cm above the floor. The simulator breathed with a sinusoidal waveform at 21.5 breaths/min with a ventilation rate of 27 l/minute. These parameters are approximately the average of the ISO standards for males and females performing moderate work.^32^ Both simulators were controlled during all experiments using custom scripted programs in LabVIEW.

### 2.3 Experimental Procedure

For experimental trials with masking conditions, a 3-ply cotton mask (Hanes Defender, HanesBrand, Inc.; Winston-Salem, NC) was fitted to the respective simulator followed by fit factor assessment using the PortaCount Pro+ (TSI, Inc.) in the N95 mode (measuring negatively charged particles 40 to 70 nm in diameter)^13^ as per manufacturer’s instructions. A daily quality assurance test was conducted using the 3M 1860 N95 respirator (Saint Paul, MN).

To test the combined effect of aerosol mitigation strategies of universal masking, physical distancing, and ventilation, experiments consisting of a matrix of the three variables were conducted (Table 1). For masking, the combinations of no masking (neither simulator wore a mask) and universal masking (both simulators wore a 3-ply cotton mask) were examined. For physical distancing, given the limitation of the distance due to the size of the environmental chamber, 0.9 m and 1.8 m distances were examined. For ventilation, four ACH rates were selected: 0, 4, 6, and 12.

**Table 1.** Experimental Parameters.

| Aerosol Generation Modality | Test Duration | Physical Distance (m) | Ventilation Rate (ACH) | Masking | Nebulizer and Simulator |
| --- | --- | --- | --- | --- | --- |
| Cough | 15 min | 0.9 and 1.8 | 0, 4, 6, 12 | No masks and universal masking | Nebulizer active during pre-cough inspiration and inactive for remainder of experiment. |
| Breathing | 15 min | 0.9 and 1.8 | 0, 4, 6, 12 | No masks and universal masking | Nebulizer active 10 seconds/inactive 50 seconds through duration of testing. Breathing continuous. |
| Breathing | 60 min | 1.8 | 0, 4, 12 | No masks and universal masking | Nebulizer active 10 seconds/inactive 50 seconds through duration of testing. Breathing continuous. |

ACH = Air changes per hour

After mask fitting and distance configuration, the environmental chamber was sealed, and the HEPA filtration system run at maximal rate to minimize background airborne particles. Thereafter, the HEPA filtration system was either turned off (0 ACH) or set to the desired air exchange rate (4–12 ACH) and allowed to run for 15 minutes, during which time all OPCs were initialized to begin particle concentration data collection and the recipient simulator activated to begin breathing. After the air exchange stabilized, the source simulator was initiated to cough or breathe, and aerosol concentrations were measured for 15 minutes. The chamber was allowed to cool to 22 °C between experiments to reduce the inter-test temperature variability. Three independent experimental replicates were conducted for each unique experimental condition.

### 2.4 Data Processing and Statistical Analysis

The background aerosol concentration was determined based on the mean particle concentration during the 3 minutes prior to cough or exhalation. The bin-specific particle counts per cubic meter of air were converted to volume based on the mean bin diameter (assuming spherical particles) and then to mass concentration by multiplying by the density of KCl (1.984 g/cm^3^). The total mass concentration was calculated by summing the bin-specific mass concentrations for all size bins. The mean mass concentration was calculated as the average mass concentration over the test duration and served as the exposure metric in these simulations. OPC data were processed using the R statistical environment v. 4.0.2 (R Project for Statistical Computing; Vienna, Austria). All point estimates are presented as the arithmetic mean ± 1 standard deviation of the measured mean mass concentration.

Regression modeling was performed in R using the base linear model (*lm*) function using a logarithmic transformation of the mean mass concentration at the mouth of the breathing recipient against three predictor variables: Masking (Unmasked = 0 and Masked = 1; categorical); Distance (0.9 m = 0 and 1.8 m = 1; categorical); Theoretical ACH (0, 4, 6, and 12; continuous). Comparisons of model fit with and without interaction between ACH and distance were conducted via an analysis of variance (ANOVA) test. Unstandardized regression coefficients are presented in addition to back-transformed coefficients expressed as percent reduction in the outcome variable (mean mass concentration). Statistical significance was set at p < 0.05.

## 3 RESULTS AND DISCUSSION

### 3.1 Chamber Conditions, Ventilation, and Aerosol Characterization

Across all experiments, the mean chamber temperature was 24.1 ± 1.1 °C with a relative humidity of 26.0% ± 2.4%. Particle clearance by the ventilation system followed first-order exponential decays, with overall clearance rates 74.1% ± 4.4% of decay rates estimated by anemometer readings (Range: 73.1%–76.7%; Figure 2A). Particle decay rates throughout the chamber, as measured by the five OPCs, were largely homogeneous (Supplemental Figure S1). The experimental decay rates after single coughs were 76.1% ± 1.5% of theoretical values (Range: 74.4%–77.3%). These experimental decay rate magnitudes and variances were comparable to those obtained from particle decay testing, which suggests that the ventilation system promoted adequate air mixing to disperse cough aerosols through the chamber volume. Therefore, we presume similar air mixing within the chamber during ventilation studies for the other two modalities tested.

**Figure 2.**
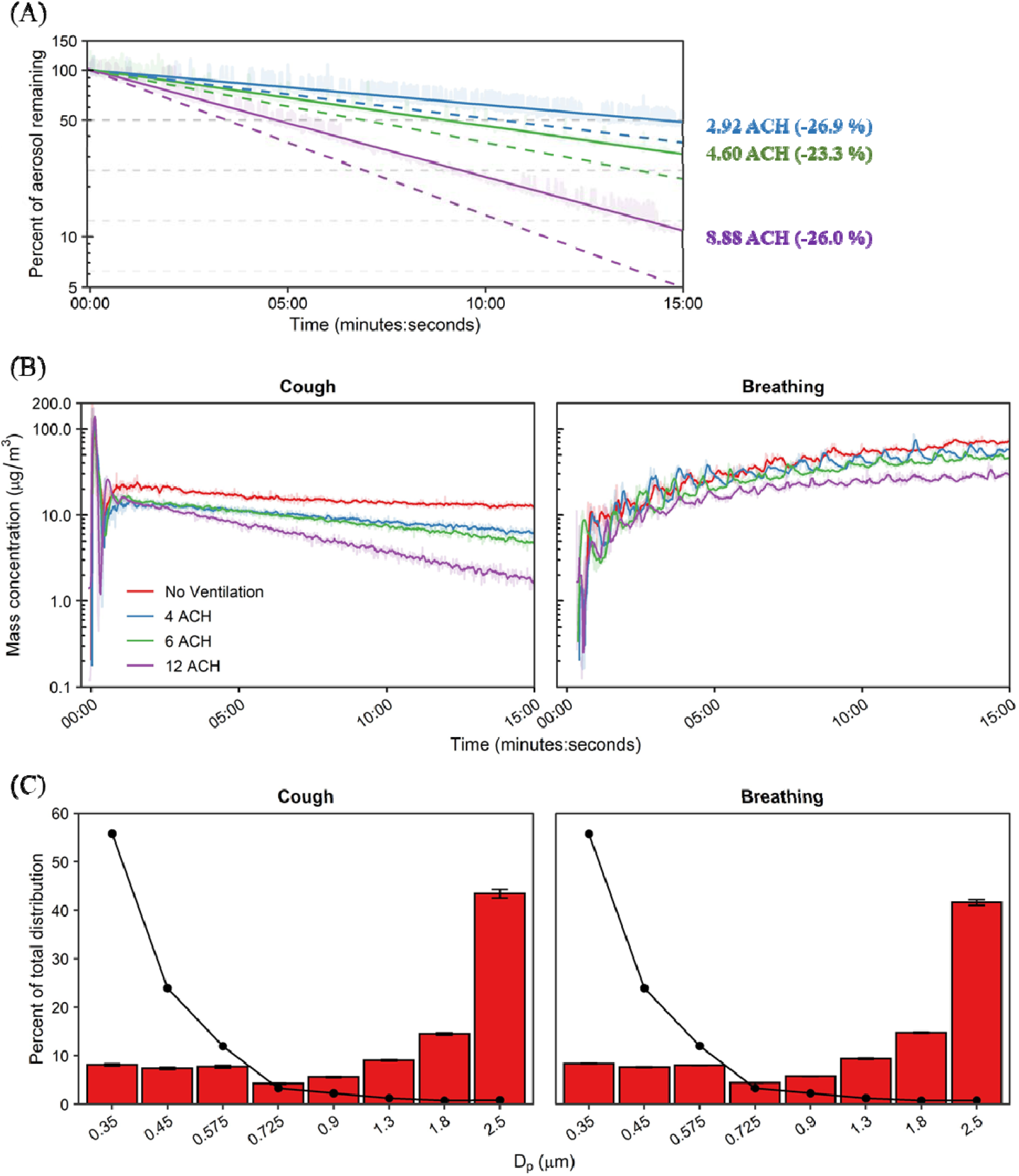
Ventilation and Aerosol Characterization. (A) Environmental chamber particle decay rates across HEPA ventilation settings. Dashed lines indicate the theoretical decay rate for each examined ventilation rate; solid lines indicate effective rates determined. Enumerated effective air exchange rates shown at the end time-point with percent error from theoretical (negative value indicates lower than theoretical). (B) Mean chamber mass concentration time curves of simulated very fine respiratory droplets and aerosol particles for the examined respiratory actions and ventilation rates. (C) Bin-specific particle distributions as determined by mass (bars) and by number of particles (line). The median particle diameter (Dp) indicates the bin. Results are the arithmetic mean ± standard deviation of three independent experiments. Error bars for the number of particles (line) too small to visualize. ACH = Air changes per hour.

Chamber aerosol concentrations during simulated respiratory events are shown in Figure 2B. A single simulated cough produced an immediate aerosol influx within the chamber followed by mixing and log-linear decay, except for no ventilation in which the aerosol concentration reached a plateau. As expected, the continuous influx of aerosol during breathing resulted in a steady increase over time, suggesting a plateau would be reached over longer testing durations in the presence of ventilation. The bin-specific size and mass distributions of the KCl aerosol averaged over the test duration were similar across modalities. The particle size data indicated that 41.2%–44.4% of the aerosol mass was in the 2–3 um range (2.5 um channel) which tapered to a nadir between 4.1% and 4.4% among the 0.65–0.8 um size range (0.725 um channel). The remaining three smallest bins each registered between 7.1% and 8.4% of the mass distribution (Figure 2C). On a particle number basis, most particles were detected within the smallest size channel with < 3% attributed to the largest size bin. Similar to the proportional mass distribution, particle count size distribution was analogous across the tested modalities. The proportional size distribution demonstrated a peak at 0.35 μm—the smallest measured size bin— via OPC, and were similar to tests measuring human respiratory aerosols via OPC-based measurements. Of note, the aerosol concentrations measured in the current investigation were 1–2 orders of magnitude higher than concentrations generated by typical human respiratory events. ^21,33-35^

### 3.2 Masking, Physical Distance, and Ventilation

The time-concentration curves at the 1.8 m physical distance are shown in Figure 3A; analogous results for the 0.9 m physical distance are presented in Supplemental Figure S2. For a single cough, aerosol concentrations decayed log-linearly after aerosol generation, while aerosol concentrations during breathing continually increased over time. Donning a 3-ply cotton mask blunted the height of the time-concentration curves. The mean aerosol mass concentrations at the mouth of the breathing recipient over the testing duration are presented in Figure 3B. Overall, universal masking reduced particle exposure compared to unmasked conditions, while exposure reduction by distance and ventilation did not produce discernable patterns of exposure modulation. To determine the relative exposure reduction of universal masking, physical distancing, and ventilation during the 15-minute tests, mean mass concentrations were regressed using ordinary least squares (OLS) multiple linear regression. Inclusion of the interaction term did not improve the model for either cough (p = 0.3062) or breathing (p = 0.6475) compared to models without interaction while also increasing the model Akaiki’s Information Criterion, thus fixed effects OLS models without interaction were constructed. Results of multiple OLS regression are presented in Table 2. Interactions between masking and the other parameters were neither expected nor tested.

**Figure 3.**
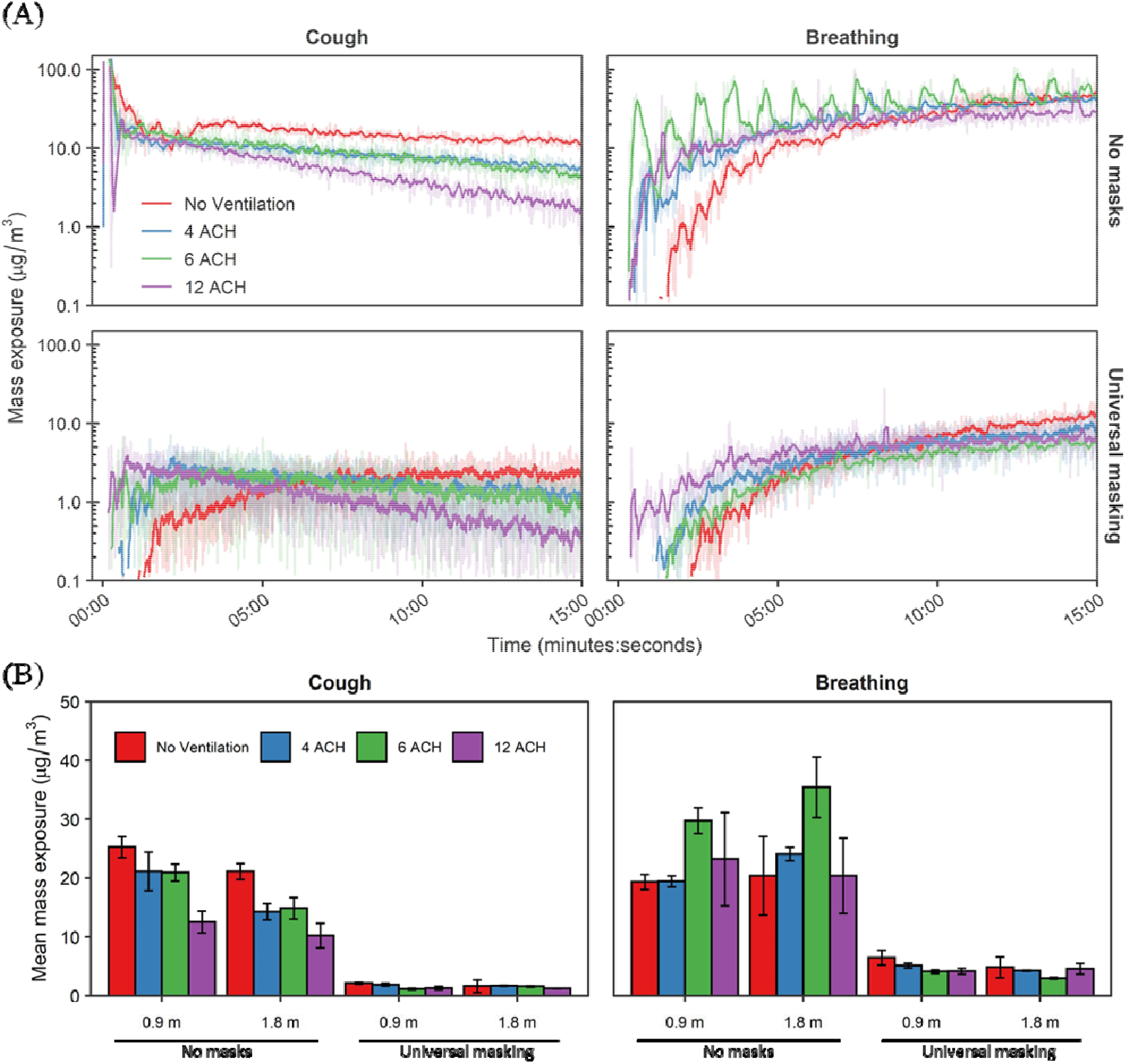
Aerosol Mass Exposure of the Recipient. (A) Mass exposure concentration over time across the matrix of modalities, masking status, and ventilation for the 1.8 m physical distance. Results for the 0.9 m physical distance are provided in the supplemental material (Supplemental Figure S2). Data are the arithmetic mean of three independent experiments. (B) Mean mass exposure over the 15-minute simulation period derived from the time curves. Results are the arithmetic mean ± standard deviation of three independent experiments. No statistical comparisons were made between individual groups. ACH = Air changes per hour.

**Table 2.**
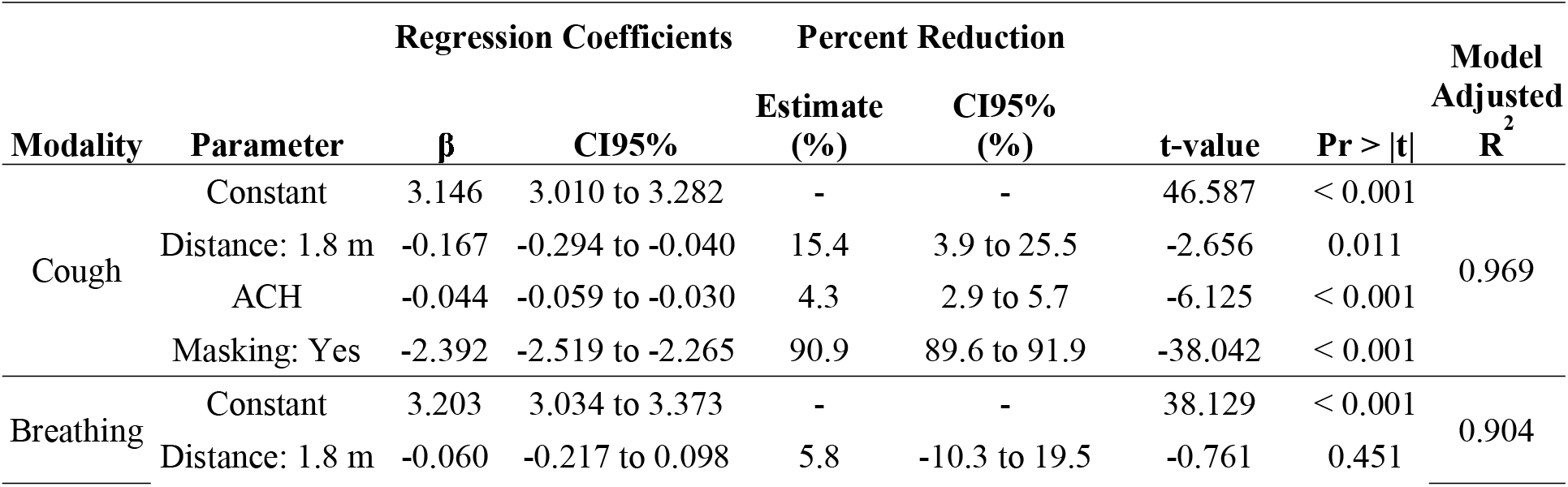
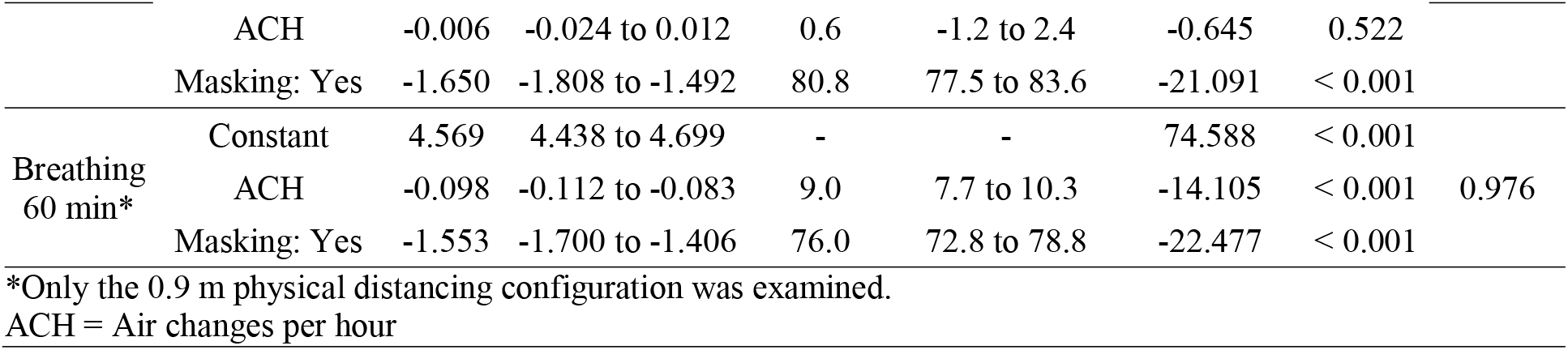
Regression Coefficients for 15- and 60-minute Tests.

Adjusting for ACH and physical distance, universal masking significantly reduced aerosol exposure compared to unmasked exposures (p < 0.001 among all modalities) during the 15-minute tests. Fit factors of the 3-ply cloth mask were 4.1 ± 2.6 (n = 43) for the recipient and 1.7 ± 0.6 (n = 42) for the source simulator. The largest reduction in aerosol mass exposure was observed after a single cough (90.9%; CI95%: 89.6%–91.9%), while exposure reduction was comparatively lower during breathing (80.8%; CI95%: 77.5%–83.6%). The reduction in mass concentration was likely due to preferential filtration of aerosols > 1 µm in diameter after having traversed two masks (Figure 4). The differences in exposure reduction among the aerosol generation modalities were likely due to specific changes in aerosol spatiotemporal dispersion when the source was masked. Aerosol plumes generated during both breathing modalities and a single cough escape through face seal leaks.^36^ The plumes would then be deflected behind and/or to the side of the source and thus effectively farther from the recipient compared with the experiments with no masks. Without chamber mixing, as observed with no ventilation, the cough aerosol deflected by the mask took longer to disperse throughout the chamber compared to without a mask as was observed in Figure 3A. The time concentration curves for breathing shifted to the right when masked, though not as much as after a single cough, showing that dispersion kinetics likely played a larger role in the heterogeneity observed for exposure reduction among the respiratory actions simulated here. While we cannot rule out the possibility that differential filtration of the source’s mask was influenced by aerosol generation (for example, the higher expulsion velocity during coughing causing greater mask aerosol filtration compared to breathing), our previous work suggests the aerosol generation modality likely does not influence mask collection efficiency for this 3-ply cotton mask (51.7% ± 7.1 % for coughing and 44.3% ± 14.0% for breathing).^13^ Lastly, we have previously shown the recipient’s breathing can contribute towards airflow patterns and subsequently influence their specific exposure^37^.

**Figure 4.**
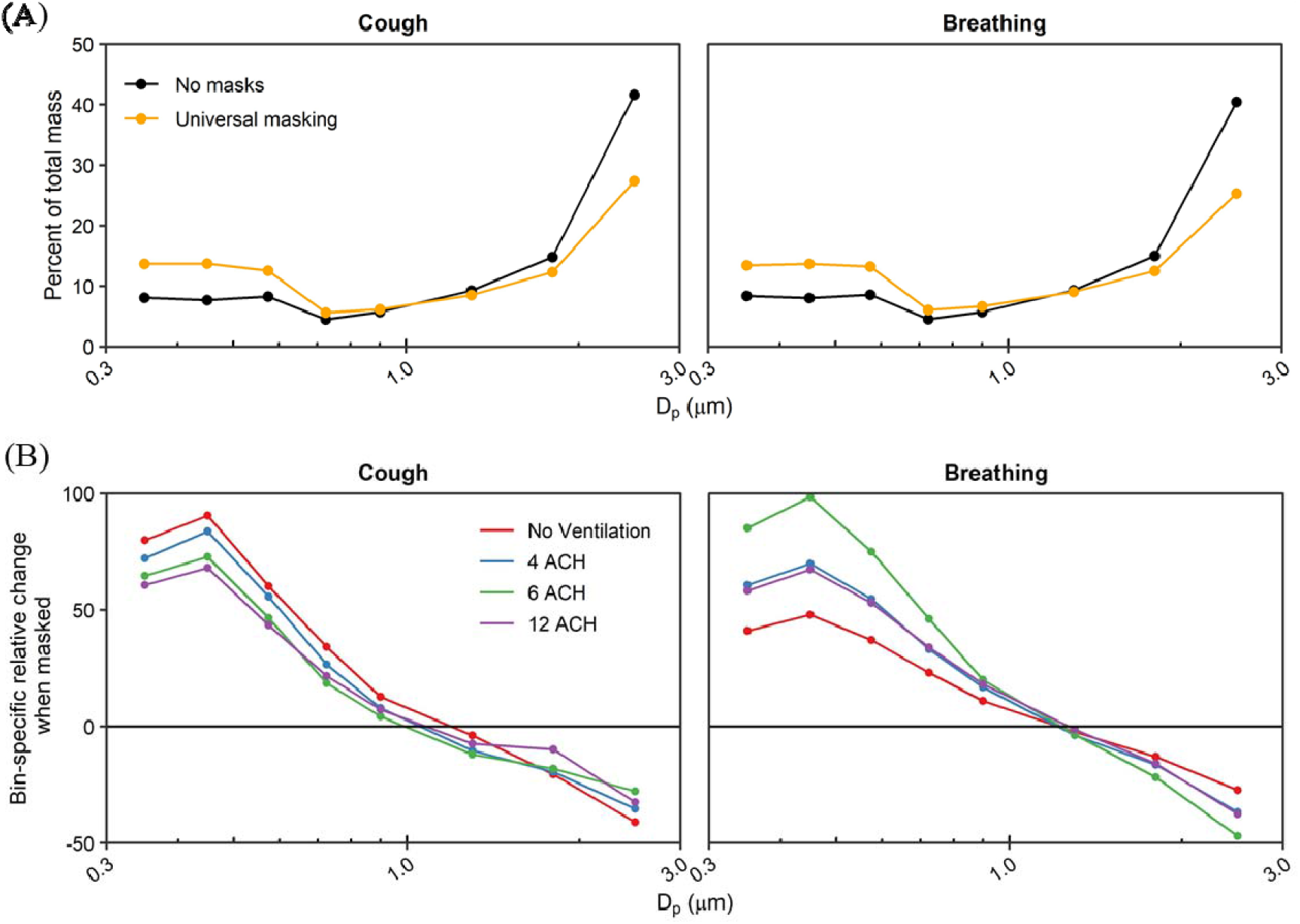
Aerosol Size Distribution Shifts during Masking. (A) Bin-specific percent of mass distribution averaged across all ventilation rates. (B) Bin-specific percent change of aerosol distribution stratified by ventilation rate. The median particle diameter (Dp) indicates the bin. Data are the arithmetic mean of three independent experiments. ACH = Air changes per hour.

The exposure reductions associated with the other predictor variables varied depending on the respiratory action for the 15-minute tests. When controlling for masking, increasing physical distance from 0.9 m to 1.8 m significantly reduced aerosol exposure from a single cough by 15.4% (CI95%: 3.9%–25.5 %; p = 0.011); increasing ventilation also reduced exposure by 4.3% per ACH (CI95%: 2.9%–5.7%; p < 0.001). Neither increasing ACH (p = 0.522) nor increasing physical distance (p = 0.451) provided protection during breathing for the 15-minute tests. When extending the test duration to 60 minutes for breathing, the mean mass concentration from aerosol generation reached a dynamic equilibrium with each of the examined ACH rates (Figure 5). Analysis for the 60-minute tests by OLS regression demonstrated increasing ventilation significantly decreased mean mass concentration by 9.0% (CI95%: 7.7%–10.3%; p < 0.001; Table 2), while universal masking expectedly reduced mean mass concentration significantly. When condensing the total aerosol generation period to the initial 3 minutes in the short-term aerosol generation tests, increasing ACH became a significant predictor in exposure reduction (5.2%; CI95%: 3.8%–6.5%; p < 0.001; supplemental table 2). The time-concentration curves of the short-term aerosol generation tests demonstrated the log-linear decay similarly to time-concentration profiles observed from a single cough, albeit shifted to the right to reflect the longer aerosol generation period (Supplemental Figure S3). This result demonstrates that attainment of a dynamic equilibrium with continuous aerosol input or removal of aerosols produced by an intense, short-term generation events through increasing ventilation can result in significant exposure reduction for a recipient. We did not examine the extended exposure duration for a single cough over 60 minutes, though we expect increasing ventilation will remain a significant predictor of mean mass concentration reduction.

**Figure 5.**
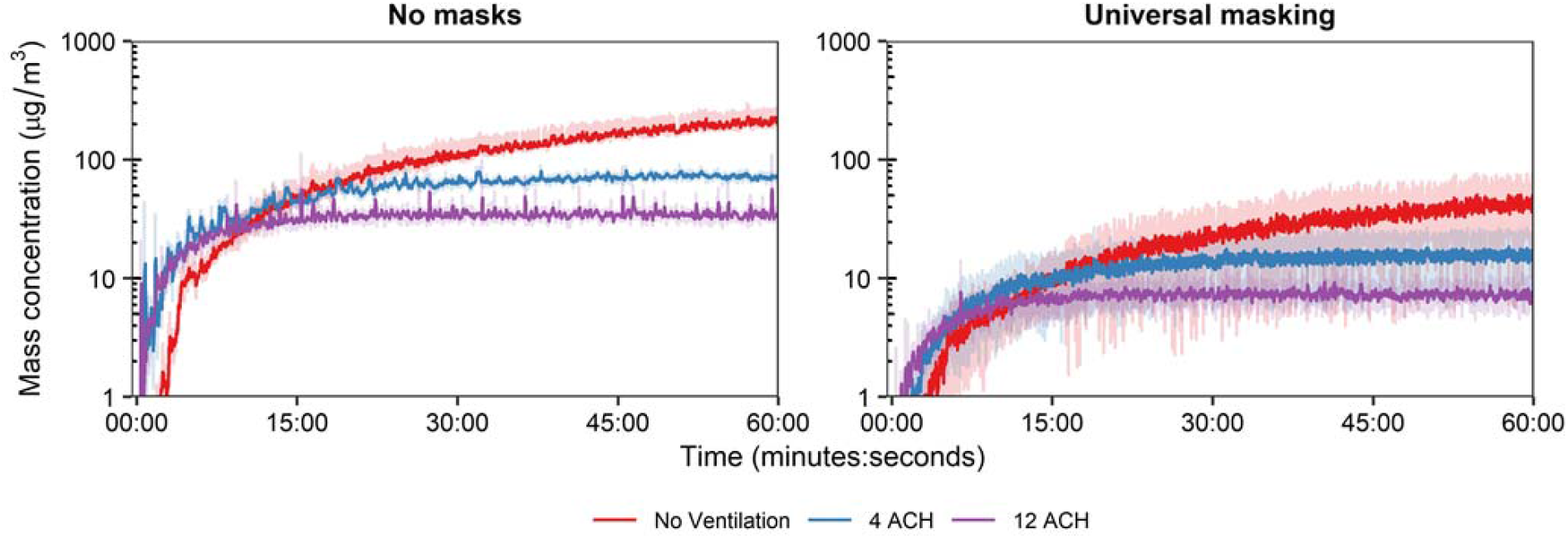
Extended Breathing Assessment. Mass exposure concentration time curves over 60-minutes for breathing at the 1.8 m physical distance and three ventilation rates. Results are the arithmetic mean ± standard deviation of three independent experiments. ACH = Air changes per hour.

With respect to ventilation, the restricted 15-minute exposure duration contributed to the lack of pronounced effect of increasing ACH for breathing and can be explained when considering air flow. Ventilation not only provides contaminant removal, but also impacts the overall air flow. Modeling of aerosol dispersion through central ventilation systems demonstrates this complex interplay between ventilatory clearance and overall air flow patterns that can, under certain situations, increase the short-term exposure during rapid, thorough mixing^38^ that was observed during the breathing respiratory action. Increasing ventilation reduced monotonically the bulk aerosol concentration throughout the entire chamber over the total duration of the ventilation testing (Figure 3A) but tended to decrease the time of aerosol contact at the mouth of the recipient. Therefore, ventilation tended to increase the recipient aerosol exposure at the onset of testing where, in extreme cases, ventilation paradoxically increased the mean mass exposure (Figure 3B); this observation was independent of physical distance. The authors opine such increases were caused by the observed thorough air mixing, as was noted in particle decay studies, in conjunction with the short exposure duration of 15 minutes. As previously noted, the effect of ventilation was appreciated during the 60-minute breathing test and the short-term aerosol generation tests. These results demonstrate that the aerosol reduction measures by ventilation must consider the air mixing, aerosol spatial dispersion, and exposure duration in addition to other mitigation strategies to ascribe the degree of protection afforded. This becomes evident when examining the physical distancing within a well-mixed environment: the effectiveness of physical distancing can diminish.^31,39,40^ Indeed, we observed apparent reduction in time to first contact with increasing ventilation at 0.9 m physical distancing (Figure S2) that was similar to the 1.8 m results. These results demonstrate that a complex interplay between air mixing and exposure duration can determine an individual’s aerosol exposure.

### 3.3 Limitations

The current investigation has several noteworthy limitations that must be considered. First, the mass concentration of aerosol generated during the experimental modalities, particularly breathing, was higher than those produced from human exhalations.^21^ The higher concentrations combined with the wide dynamic range of the OPC allowed for stable and reproducible measurements while assuring attainment of quantitative limits of detection among all tests. Second, the simulators lack generation of body heat and the ability to generate a thermal exhalation plume, which affects aerosol dispersion and inhalation exposure.^41^ Given the confines of the environmental chamber, the internal ventilation setup, and the high aerosol concentrations, we would not expect substantial differences in mean mass exposure given the small volume of the chamber. Therefore, limits must be placed on the interpretation of the results within a larger indoor environment, especially considering the dispersion potential of an exhalatory thermal plume. Third, the range of human respiratory aerosols can be smaller and larger than the measured range of this investigation (0.3–3.0 µm).^33,35^ For droplets, the effect of physical distancing may be higher than those suggested by the observed results for droplets. Fourth, the study investigated the exposure reduction of a single 3-ply cotton mask. The authors recognize the limitation of having tested a single mask, since the effectiveness of exposure reduction by other masks could be either higher or lower, depending on the mask. Nonetheless, the analytics of the study allow for reasonable expectation of exposure reduction of the other predictor variables provided the aerosol behavior does not significantly deviate from this study with another type of mask.

## 4 CONCLUSION

The current investigation highlights the contribution of three common engineering and administrative controls recommended for limiting SARS-CoV-2 exposure within an indoor environment: ventilation, physical distancing, and universal masking. When controlling for the other two mitigation strategies, universal masking with a 3-ply cotton mask contributed to the plurality of the observed reduction in aerosol mass exposure irrespective of aerosol generation modality. This reduction was due, in part, to preferential reduction of particles > 1.0 μm in diameter from reaching the recipient. The overall effectiveness of ventilation and physical distancing was dependent upon the modality under simulated conditions. Although masking alone provided good exposure reduction, a combination of engineering and administrative controls remains important in reducing an individual’s personal exposure to potentially infectious very fine respiratory droplets and aerosol particles within an enclosed indoor space.

## Data Availability

The datasets generated during and/or analyzed during the current study are available from the corresponding author on reasonable request.

## ACKNOWLEDGEMENTS

The authors wish to acknowledge the facilities, maintenance, and security personnel of NIOSH Morgantown for their hard work and dedication, which were integral to the completion of this work.

## DISCLAIMER

The findings and conclusions in this report are those of the authors and do not necessarily represent the official position of the National Institute for Occupational Safety and Health, Centers for Disease Control and Prevention.

