## Supplemental File 1 for "Reduction of exposure to simulated respiratory aerosols using ventilation, physical distancing, and universal masking"

Dr. William G. Lindsley

National Institute for Occupational Safety and Health (NIOSH)

1000 Frederick Lane, M/S 4020

Morgantown, WV 26508-5402

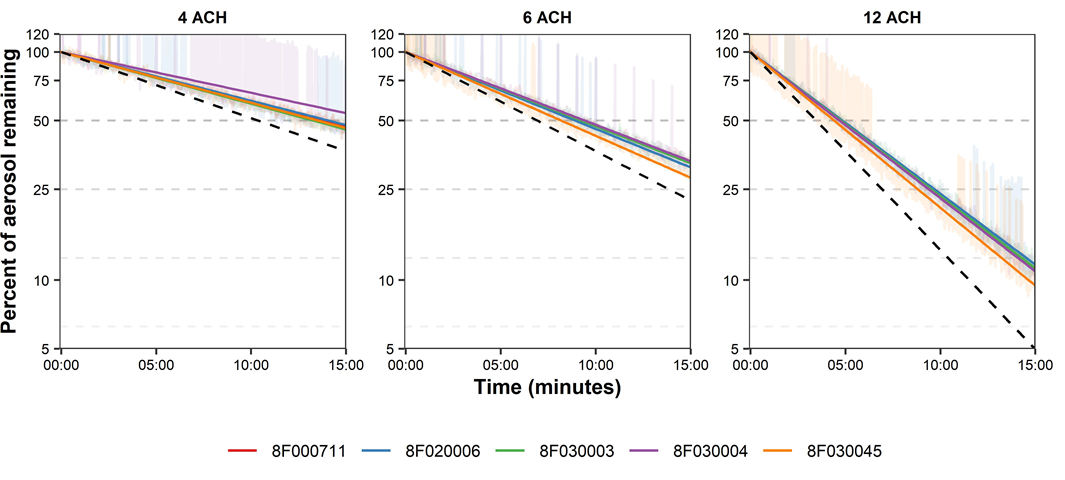

Supplemental Figure S1. Chamber Aerosol Decay Curves for each Grimm Optical Particle Counter. NaCl particles were produced with the TSI Particle Generator after particles from the smallest three bins were measured in real-time for 15 minutes at the examined ventilation rates. The exponential decay curves are overlain (solid lines) with theoretical decay rates (dotted lines) for a well-mixed room. ACH = Air changes per hour.

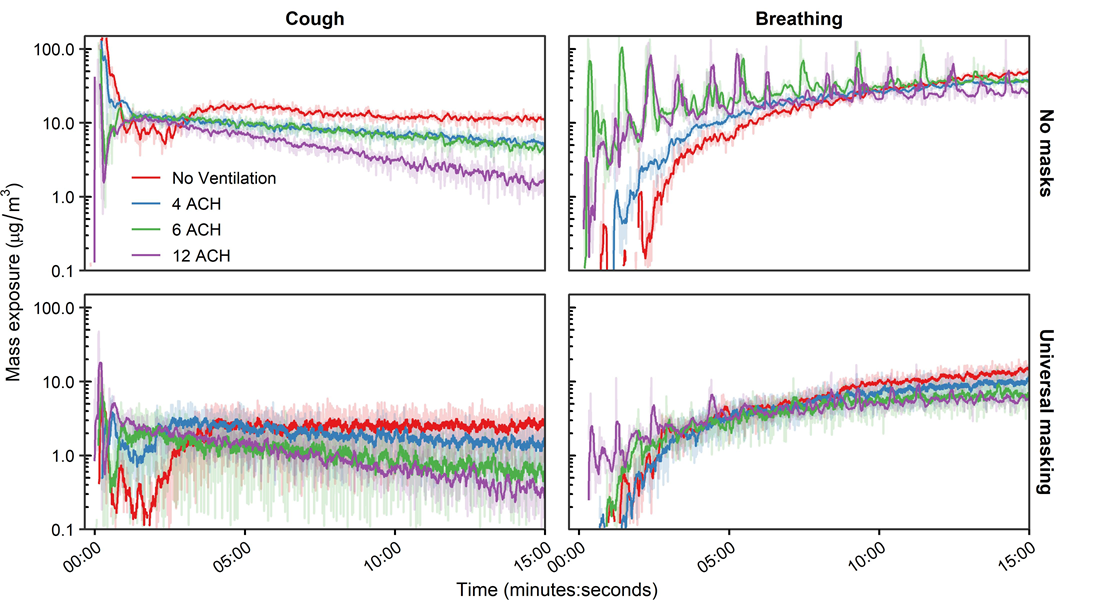
Supplemental Figure S2. Aerosol Mass Exposure of the Recipient. Mass exposure concentration time curves for a single cough and breathing, masking status, and ventilation for the 0.9 m physical distance. Results are the arithmetic mean of three independent experiments. ACH = Air changes per hour.

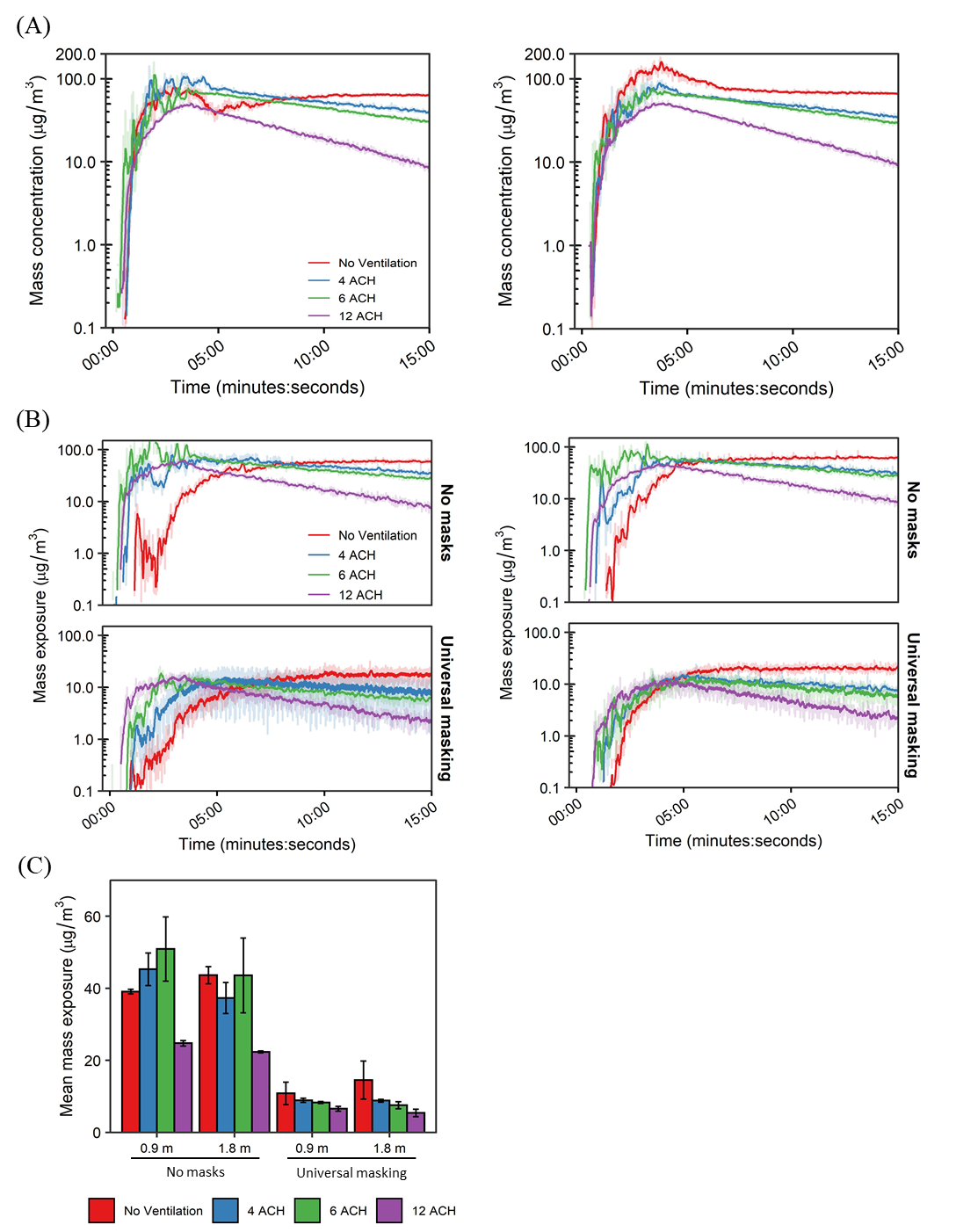

Supplemental Figure S3. Aerosol Mass Exposure of the Recipient in Short-term Aerosol Generation Tests. A) Aera sampler time-concentration curves for the 0.9 m (Left) and 1.8 m (Right) physical distance. B) Time-concentration curves of aerosol exposure at the mouth of the recipient across the masking and ventilation. The curves for the 0.9 m (Left) and 1.8 m (Right) physical distance are presented for comparison. C) Mean mass exposure over the 15-minute simulation period derived from the time curves. Results are the arithmetic mean ± standard deviation of three independent experiments. ACH = Air changes per hour.

| **Supplemental Table 1. Regression Coefficients for Short-term Aerosol Generation Test** | | | | | | | | |
| --- | --- | --- | --- | --- | --- | --- | --- | --- |
| **Modality** | **Parameter** | **Regression Coefficients** | | **Percent Reduction** | | **t-value** | **Pr > \|t\|** | **Model Adjusted R^2^** |
|  |  | **β** | **CI95%** | **Estimate (%)** | **CI95%**  **(%)** |  |  |  |
| Short-term Aerosol Generation | Constant | 3.923 | 3.792 to 4.054 | - | - | 60.394 | < 0.001 | 0.933 |
|  | Distance: 1.8 m | -0.050 | -0.172 to 0.072 | 4.9 | -7.4 to 15.8 | -0.829 | 0.412 |  |
|  | ACH | -0.053 | -0.067 to -0.039 | 5.2 | 3.8 to 6.5 | -7.577 | < 0.001 |  |
|  | Masking: Yes | -1.477 | -1.599 to -1.355 | 77.2 | 74.2 to 79.8 | -24.423 | < 0.001 |  |
| ACH = Air changes per hour | | | | | | | | |
